# ROBI: a Robust and Optimized Biomarker Identifier to increase the likelihood of discovering relevant radiomic features

**DOI:** 10.1101/2024.09.09.24313059

**Authors:** Louis Rebaud, Nicolò Capobianco, Clémentine Sarkozy, Anne-Ségolène Cottereau, Laetitia Vercellino, Olivier Casasnovas, Catherine Thieblemont, Bruce Spottiswoode, Irène Buvat

## Abstract

**Objectives:** The Robust and Optimized Biomarker Identifier (ROBI) feature selection pipeline is introduced to improve the identification of informative biomarkers coding information not already captured by existing features. It aims to accurately maximize the number of discoveries while minimizing and estimating the number of false positives (FP) with an adjustable selection stringency.

**Methods:** 500 synthetic datasets and retrospective data of 378 Diffuse Large B Cell Lymphoma (DLBCL) patients were used for validation. On the DLBCL data, two established radiomic biomarkers, TMTV and Dmax, were measured from the 18F-FDG PET/CT scans, and 10,000 random ones were generated. Selection was performed and verified on each dataset. The efficacy of ROBI has been compared to methods controlling for multiple testing and a Cox model with Elasticnet penalty.

**Results:** On synthetic datasets, ROBI selected significantly more true positives (TP) than FP (p < 0.001), and for 99.3% of datasets, the number of FP was within the estimated 95% confidence interval. ROBI significantly increased the number of TP compared to usual feature selection methods (p < 0.001). On retrospective data, ROBI selected the two established biomarkers and one random biomarker and estimated 95% chance of selecting 0 or 1 FP and a probability of 0.0014 of selecting only FP. Bonferroni correction selected no feature, and Elasticnet selected 101 spurious features and discarded TMTV.

**Conclusion:** ROBI selected relevant biomarkers while effectively controlling for FPs, outperforming conventional selection methods. This underscores its potential as a valuable asset for biomarker discovery.

**Highlights:**

- ROBI is a feature selection tool capable of screening thousands of features.
- It enables systematic evaluation of numerous radiomic features while minimizing false discoveries.
- ROBI was validated on synthetic and real datasets, outperforming other selection techniques.

## Introduction

Radiomics involves the extraction and analysis of quantitative medical image features (1,2). By converting images into mineable data, radiomics may reveal disease characteristics that are currently overlooked, improving diagnosis, prognosis, and treatment planning. A great number of scientific publications have mentioned radiomics since its introduction, but reproducibility, standardization, interpretability, and methodological issues limit its potential, and few radiomics results have been translated into the clinic (3,4).

Standards for radiomic feature definition and calculation, and guidelines for best practices are being developed to accelerate clinical translation (5-8). Lack of external validation and methodological flaws in assessing biomarker novelty and prognostic power partly explain why radiomics has not been adopted in the clinics yet. Statistical methods, such as robust feature selection algorithms, cross-validation techniques for model evaluation, and statistical tests for assessing the significance of prognostic biomarkers, can address some of these challenges by ensuring the reliability and generalizability of radiomics studies. On the other hand, improper use of these techniques—including overfitting models to specific datasets, not controlling for C-index inflation, ignoring multiple testing corrections, data leakage in the machine learning pipeline and failing to validate findings externally—can lead to misleading results, characterized by either overly optimistic or pessimistic evaluations of radiomic features and models.

In this context, we introduce the Robust and Optimized Biomarker Identifier (ROBI), not as a novel feature selection method, but as a software solution designed to combine a range of established techniques in a simple yet efficient manner. ROBI is a streamlined Python package designed to facilitate the selection of radiomic features, thereby mitigating the risk of selecting features that either mirror existing biomarkers (Orlhac et al., 2014 (9)) or lack prognostic relevance. By implementing current best practices within an optimized framework, ROBI aims to minimize false positives—erroneously selected non-relevant features—while enhancing the detection of true positives—genuinely relevant features. It employs time-efficient permutation tests to precisely estimate the number of false positives, offering users the flexibility to tailor selection stringency according to their research objectives.

ROBI’s efficacy is demonstrated through validation on synthetic datasets with established truths, and on a cohort of Diffuse Large B Cell Lymphoma (DLBCL) patients, where it successfully identified two known biomarkers out of many random ones. This underscores ROBI’s utility as a practical tool that leverages existing methodologies to overcome some of the current barriers in radiomics, paving the way for more reliable and clinically applicable radiomic research outcomes.

## Material and methods

### 1. Pipeline

Candidate biomarkers (CB) are assessed for their predictive potential by ROBI, based on their values in a patient cohort and their association with the outcome (e.g., time before relapse, response to treatment). To avoid selecting candidates that replicate known predictive information, previously known predictive biomarkers must be identified. Figure 1 presents the overall pipeline. More details on the choice of the parameter values are provided in the supplemental data.

**Figure 1:**
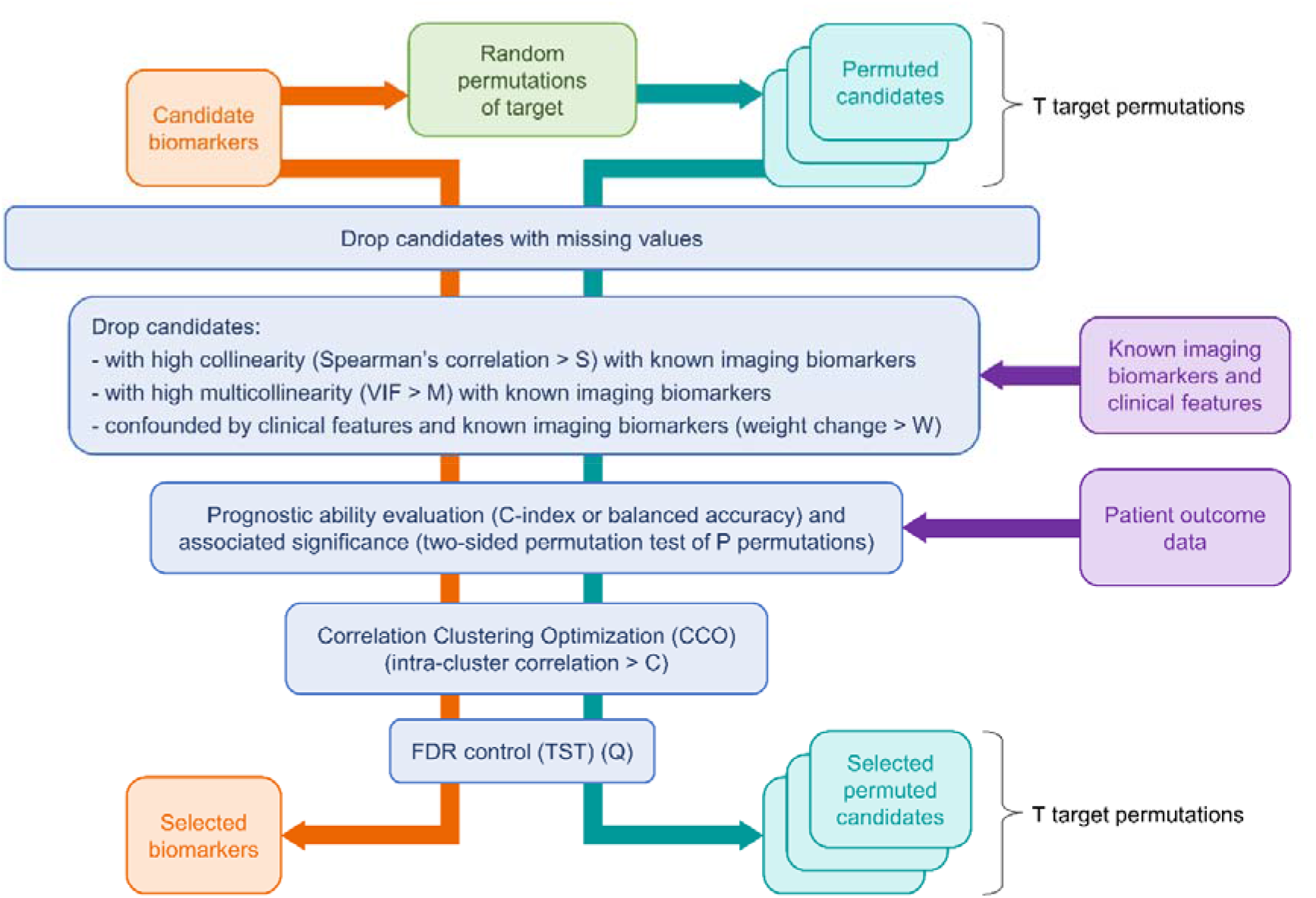
Diagram of the ROBI selection pipeline. Each free tuning parameter is denoted by a capital letter (S, M, W, P, C, Q and T). Intuitive explanation and range of values of these parameters are provided in supplemental materials. “VIF” is the Variance Inflation Factor. “weight change” is the relative change in weight when confounders are introduced. “FDR” is the False Discovery Rate and “TST” stands for two-stage linear step-up procedure, the technique used to control for FDR. Filtering candidates reproducing known information and CCO are optional.

#### a. Discarding missing values

A low number of samples and high numbers of censored samples artificially increase biomarker prognostic value (10,11). Any CB with missing values is thus discarded to avoid favoring CBs unavailable to all patients.

#### b. Discarding already known information

CBs with an absolute Spearman correlation coefficient greater than a tunable cut-off S (0.5 by default) with a known imaging or clinical biomarker are discarded to ensure that the selected CB capture new information. In case of multiple established biomarkers, multicollinearity is assessed using the Variance Inflation Factor (VIF). CBs exceeding a certain tunable multicollinearity threshold M (5 by default) are discarded. A linear model (Cox for survival and logistic regression for classification) controls for confounders (e.g., age, treatment) (12). A univariate model with only the evaluated CB is trained first and assigns a weight W_*uni*_ to the CB. Then, a multivariate model with the evaluated CB and known covariates is trained and the new weight W_*multi*_ is assigned to the CB. The relative change in weight is defined as:

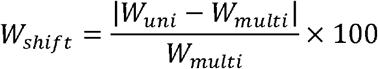

Any CB with W_*shift*_ above a threshold W (10% by default) is discarded.

#### c. Assessment of CBs performance

Each CB’s prognostic ability is assessed using Harrell’s Concordance Index (C-index) against patient outcome data such as time of death or relapse, accommodating censored outcomes, or balanced accuracy for classification task, accommodating imbalanced datasets. These scores are tested for significance using a two-sided permutation test of P permutations (1,000 by default). A two-stage linear step-up procedure (TST) is used to control the false discovery rate (FDR, the proportion of false positive in selected biomarkers) and address multiple testing (13). This statistical method uses a conservative threshold to identify potential selection and adjusts this threshold in a second stage based on the initial results to increase power while controlling the overall FDR. Adjusting TST’s Q parameter allows flexibility in balancing numbers of FPs and selected CBs. To increase the yield, ROBI performs the TST last in the selection process when the number of tested CBs has already been substantially reduced through the previous selection steps.

#### d. Optimization of the number of selected biomarkers

To optimize the selection of biomarkers, we employ a correlation clustering optimization (CCO) strategy, where CBs conveying similar information are grouped based on their absolute Spearman’s correlation. Within each cluster, only the biomarker demonstrating the highest predictive accuracy is retained. This approach is informed by methods previously utilized in genomics, notably the weighted gene correlation network analysis (WGCNA) technique, which clusters genes based on similarity in expression patterns to identify modules of highly correlated genes, thereby facilitating the interpretation of complex biological phenomena (14). By adopting a similar methodology, we adjust the maximum allowable correlation between two clusters, C (0.5 by default), to fine-tune the granularity of the clustering and thus the number of biomarkers selected. This method not only enhances the specificity of our biomarker selection process but also ensures that the biomarkers retained offer of more unique predictive value, thereby avoiding redundancy.

#### e. False positive estimation

Because it is selecting the CBs with the best p-values, CCO may optimistically bias TST’s FDR. To correct and improve the number of FP estimation, ROBI randomly permutes outcome data during selection. This preserves the relationships among CBs but breaks their association with patient outcomes. The features selected using the permuted outcome are thus FP. After repeating this process T times (by default 1,000 times), ROBI calculates the average number of FPs and its 95% confidence interval. The probability of only selecting FPs is assessed by the proportion of permutated datasets with as many as or more selected CBs than the non-permuted selection.

### 2. Synthetic data evaluation

A total of 500 synthetic datasets were generated with scikit-learn (15) and scikit-survival (16) Python packages to evaluate ROBI. These datasets varied in the number of samples, number of genuine (associated with the outcome) and spurious (not associated with the outcome) biomarkers, censoring, correlation between biomarkers, and target noise. Table 1 shows the parameter distributions and ranges. Details about the generation of the datasets are provided in the documentation of scikit-learn (15).

**Table 1:**
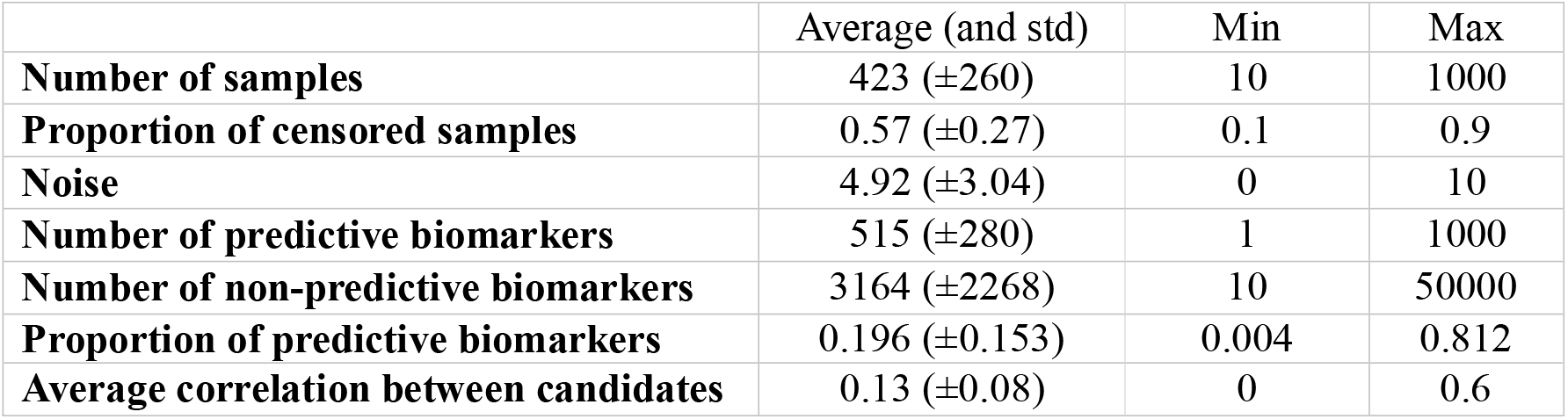
Average, standard deviations, and range of the synthetic dataset features.

A linear regression with random weights on genuine biomarkers defined the target using Scikit-learn’s “make_regression” function. The target was not built using spurious biomarkers. ROBI processed each dataset with CCO with P = 10^6^, and T = 10^3^. Genuine biomarkers selected by ROBI were defined as TPs and selected spurious biomarker as FPs. The “effective_rank” parameter within “make_regression” allowed for the simulation of correlations among features (biomarkers) by controlling the rank of the covariance matrix used to generate the features. A lower “effective_rank” implies a higher correlation among a subset of features, thereby simulating real-world scenarios where biomarkers might exhibit interdependencies. The same datasets were also processed with the two-stage linear step-up procedure (TST) alone to compare its results to ROBI’s selections and verify that ROBI’s optimization improves the number of biomarkers rightly selected.

The average, standard deviation and 95% confidence interval of the number of selected CBs, TPs and FPs were calculated as well as the percentage of datasets with more TPs than FPs, for different values of Q. Wilcoxon signed-rank tests were used to determine whether ROBI selected more TPs than FPs and if using ROBI increased the number of rightly selected biomarkers compared to TST alone. The distribution of the difference of TPs for the ROBI and TST selection for the same number of FPs was plotted.

### 3. Real data evaluation

DLBCL patients from REMARC (NCT01122472) and LNH073B (NCT00498043) cohorts were analyzed. Detailed cohort compositions have been described elsewhere (17,18). All patients had baseline anonymized ^18^F-FDG PET/CT scans, Progression Free Survival (PFS) and Overall Survival (OS) available. Lesions were segmented by expert nuclear medicine physicians (ASC, LV, MM) in the PET images (19,20).

In DLBCL, Total Metabolic Tumor Volume (TMTV) and maximum distance between two lesions (Dmax) are known to be prognostic of PFS and OS (20,21). These two biomarkers were calculated on the segmented PET images using PyRadiomics (21)

10,000 spurious biomarkers were randomly generated for all patients and input to ROBI in addition to TMTV and Dmax. ROBI parameter settings were S = 0.5, M = 5, W = 10%, P = 10^7^ and T = 10^4^. Q was set to have at least one selected CB. No CCO was used because spurious biomarkers are random and have low correlation. Biomarkers were tested to not replicate the information of ECOG, age adjusted International Prognostic Index (23), treatment, and sex. We then checked whether TMTV and Dmax were selected by ROBI and whether the number of selected spurious biomarkers was within the 95% confidence interval of ROBI’s estimated number of FPs. Selection was performed for progression (PFS) or death from any cause (OS) prediction.

Selection was also performed with other feature selection techniques: TST with a Q value chosen to have less than one false positive, the Bonferroni procedure with a probability of 0.05 of having 1 or more false positive, and a Cox model with Elasticnet penalty.

## Results

### 1. Synthetic data evaluation

A total of 99.3% of datasets had the number of FPs within ROBI’s 95% confidence interval. Table 2 shows ROBI’s selection results on synthetic datasets, and Figure 2 shows the average number of selected features and FPs, with their 95% confidence intervals, as a function of Q, for both ROBI’s and TST’s selection. More CBs were selected with higher Q. ROBI selected significantly more TPs than FPs (p < 0.001). For the same Q, ROBI significantly increased numbers of TPs, FPs, and the difference between them compared to TST (p < 0.001).

**Table 2:** Average values and standard deviation of the number of selected candidate biomarkers (CB), number of true positives (TP), false positives (FP), and percentage of datasets with more FP than TP, for different levels of Q, for the ROBI pipeline and the TST procedure alone.

| **Q** | **Method** | **Number of selected CB** | **Number of TP** | **Number of FP** | **Percentage of datasets with more TP than FP** |
| --- | --- | --- | --- | --- | --- |
| **0.01** | TST | 4 ($\pm 20$) | 4 ($\pm 20$) | 0 ($\pm 0$) | 100.0 % |
| | ROBI | 6 ($\pm 28$) | 6 ($\pm 27$) | 0 ($\pm 0$) | 99.8 % |
| **0.10** | TST | 9 ($\pm 33$) | 9 ($\pm 32$) | 0 ($\pm 0$) | 99.8 % |
| | ROBI | 15 ($\pm 47$) | 14 ($\pm 44$) | 1 ($\pm 12$) | 99.2 % |
| **0.25** | TST | 13 ($\pm 43$) | 13 ($\pm 42$) | 0 ($\pm 2$) | 99.4 % |
| | ROBI | 28 ($\pm 81$) | 21 ($\pm 56$) | 6 ($\pm 48$) | 97.1 % |
| **0.50** | TST | 19 ($\pm 57$) | 17 ($\pm 52$) | 1 ($\pm 10$) | 99.2 % |
| | ROBI | 52 ($\pm 141$) | 32 ($\pm 71$) | 20 ($\pm 99$) | 94.6 % |

**Figure 2:**
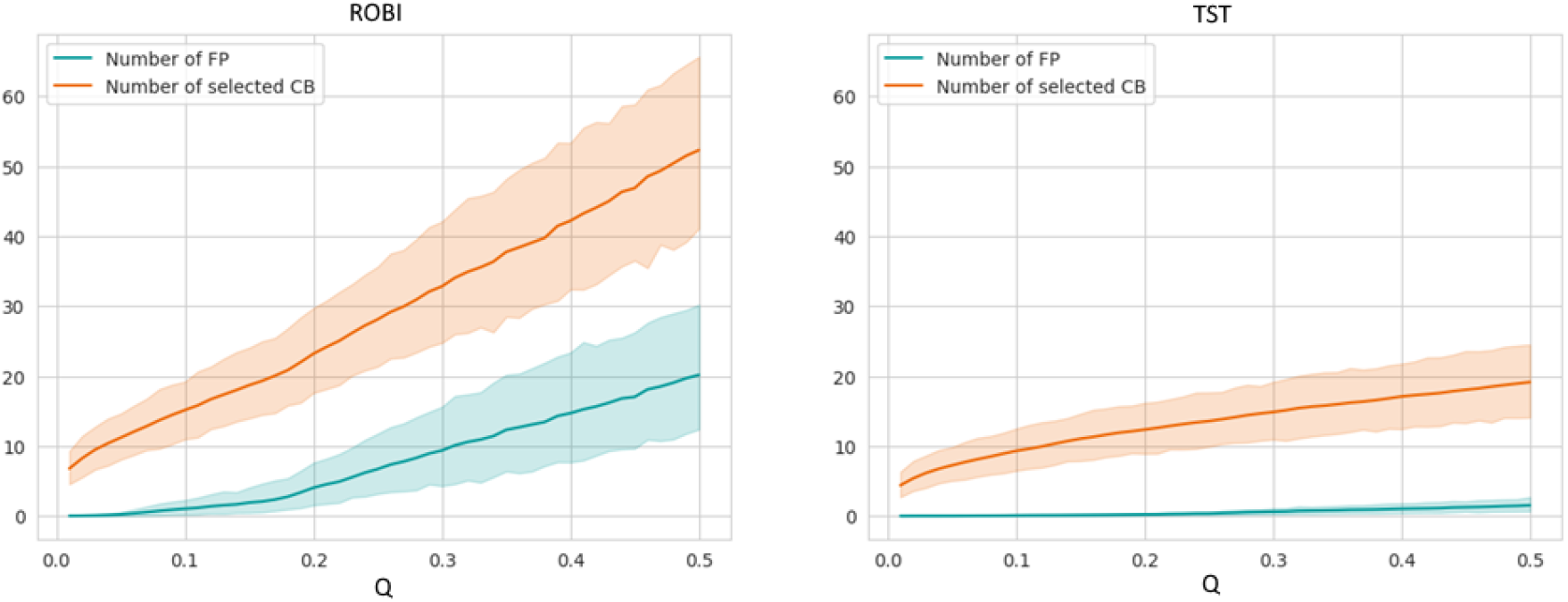
average number of selected candidate biomarkers (CB) and average number of false positives (FP) among the selected CB, with the associated 95% confidence interval, for the ROBI pipeline and the TST procedure alone, at various levels of Q.

Figure 3 plots the difference between numbers of TPs of ROBI and the number of TPs of TST for samples in which the same numbers of FPs were selected. For the same number of FPs, ROBI selected significantly more TPs than TST alone (p < 0.001).

**Figure 3:**
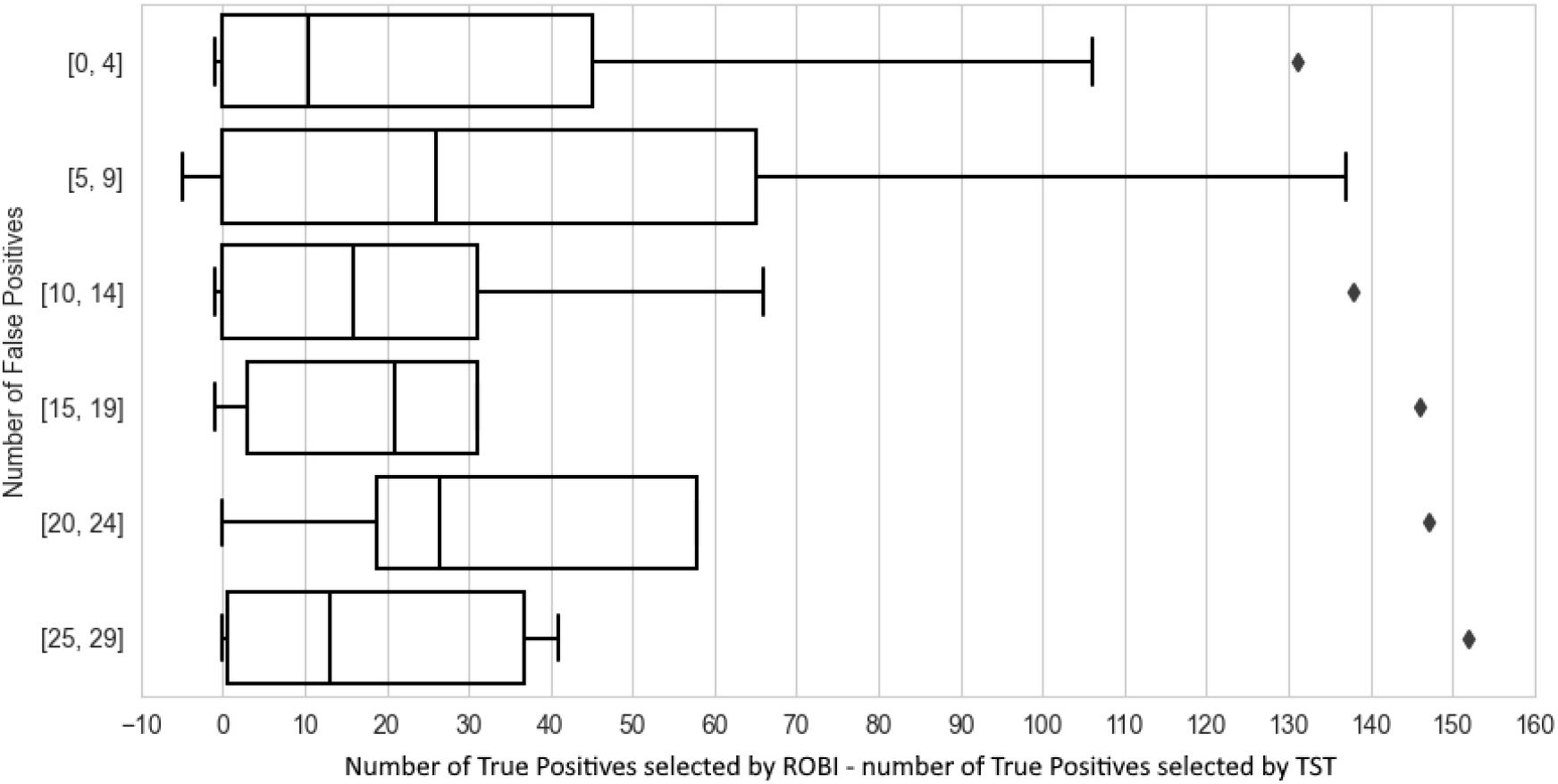
Difference between the number of True Positives (TP) selected by ROBI and the number of TPs selected by TST alone when the two methods had the same number of False Positives (FP). The difference is positive most of the time, meaning that ROBI effectively improved the rate of TP selection.

The probability of having only FPs in the selection estimated by ROBI was strongly correlated with the number of TPs (ρ = -0.96, p < 0.001). For 60% of cases with at least one TP, this probability was below 0.05. For the cases with only FPs selected (3.3% of all cases), 0.6% of them had a probability below 0.05.

### 2. Real data evaluation

The DLBCL cohort included 378 patients, among whom 96 had progressive disease and 55 died.

For PFS prediction, TMTV and Dmax both yielded a C-index of 0.63, while 105 spurious features had a C-index > 0.58 and 16 had a C-index > 0.60. The significance of the spurious features was p < 0.01 for 103 of them, and p < 0.001 for 13 of them. The Bonferroni selection did not select any feature. TST selected both TMTV and Dmax and one spurious feature. An Elasticnet selected Dmax and ranked it first, but it did not select TMTV and selected 101 spurious features. ROBI selected TMTV and Dmax, and one spurious feature. ROBI predicted a 95% chance of having 0 or 1 FP with an average of 0.1 FP. ROBI estimated the probability of having only FP to be 0.0014.

For OS prediction, TMTV and Dmax had respectively a C-index of 0.63 and 0.60, and 137 spurious features had a C-index > 0.60. The significance of the spurious features was p < 0.01 for 110 features, and p < 0.001 for 8 of them. The Bonferroni selection did not select any feature. TST did not select any spurious features, nor TMTV nor Dmax. An Elasticnet selected TMTV and ranked it 47. It did not select Dmax and selected 73 spurious features. ROBI did not select any feature.

## Discussion and Conclusion

This study introduced ROBI, the Robust and Optimized Biomarker Identifier. We called it “Robust” because false discoveries are controlled and “Optimized” because multiple strategies increase the number of rightly selected biomarkers. We showed that this selection tool efficiently controls the false positive numbers while increasing the number of selected biomarkers compared to the standard two-stage linear step-up procedure (TST) alone. ROBI’s 95% confidence interval estimating the number of false positives was correct for 99.3% of the synthetic datasets, small difference between these two numbers being probably explained by statistical fluxes. It can find relevant biomarkers among thousands of candidates with enough data (96 events for PFS prediction in our real dataset), while other standard methods fail with such a high number of potential candidates. For instance, in the test performed on the DLBCL, enough patients had PFS observed to select TMTV and Dmax, but not enough events were observed in OS to select them.

As shown by the evaluation on the synthetic datasets, ROBI’s utility transcends the radiomic domains, making it a versatile tool for biomarker selection across various fields (e.g., genomics).

ROBI has limitations. Only biomarker screening is addressed. Validating a new biomarker requires definition, measurement, standardization, modeling, and interpretation. More importantly, ROBI does not replace external validations. It only increases the chance of replicating the findings by controlling the risk of false positive selection.

Limitations include dropping candidate biomarkers with missing data. This step may eliminate promising biomarkers by reducing the number of candidates. Removing a few patients (preferably those with a censored target value) to avoid discarding too many candidate biomarkers can mitigate this limitation.

ROBI is more time consuming than other selection methods. However, thousands of biomarkers can be accurately evaluated in a reasonable time. On a PC an Intel Core i7-11800H (2.30 GHz), NVIDIA GeForce RTX 3070 (8 GB), and 16 GB RAM, 5,000 candidates could be evaluated in less than 9 min, with T = 10^3^ et P = 10^7^.

ROBI may not identify all relevant biomarkers among candidates, and the number of false negatives (predictive biomarkers that are not selected) remains unknown. In addition, because it uses univariate tests, ROBI may not always choose biomarkers that improve multivariate models. Furthermore, while the pipeline can estimate the number of false positives and the probability of selecting only false positives, it cannot tell which feature is more likely to be a true positive, and external validation remains required to validate the selected features.

Because ROBI uses a multivariate model to address confounders, only a finite number of them can be handled. For survival prediction, the general guideline is 10 non-censored samples per confounder (24).

An important future work needed is a more thorough comparison to other feature selection techniques on non-synthetic datasets.

In conclusion, ROBI selects biomarkers that best predict patient outcomes in a cohort, by discarding candidates that do not measure any new predictive information. ROBI identifies the most promising candidates, which will then have to be tested on external cohorts to confirm their predictive value. ROBI might facilitate feature selection in radiomics and beyond, and to support this effort, we provide a user-friendly Python implementation at https://github.com/Lrebaud/robi.

## Data Availability

The raw data used in this study, including PET/CT scans and clinical endpoints are not publicly available due to patient privacy requirements.

## Abbreviations

ROBI: Robust and Optimized Biomarker Identifier
FP: False Positive
TP: True Positive
CB: Candidate biomarker
TST: two-stage linear step-up procedure
FDR: False discovery rate
CCO: Correlation Clustering Optimization
DLBCL: Diffuse Large B Cell Lymphoma
TMTV: Total Metabolic Tumor Volume
Dmax: maximum distance between two lesions

## References

1. Gillies RJ, Kinahan PE, Hricak H. Radiomics: Images Are More than Pictures, They Are Data. Radiology. 2016;278:563–577. doi: 10.1148/radiol.2015151169

2. Song J, Yin Y, Wang H, Chang Z, Liu Z, Cui L. A review of original articles published in the emerging field of radiomics. Eur J Radiol. 2020;127:108991. doi: 10.1016/j.ejrad.2020.108991

3. Volpe S, Mastroleo F, Krengli M, Jereczek-Fossa BA. Quo vadis Radiomics? Bibliometric analysis of 10-year Radiomics journey. Eur Radiol. 2023;33:6736–6745. doi: 10.1007/s00330-023-09645-6

4. Pinto dos Santos D, Dietzel M, Baessler B. A decade of radiomics research: are images really data or just patterns in the noise? Eur Radiol. 2021;31:1–4. doi: 10.1007/s00330-020-07108-w

5. Koçak B, Durmaz EŞ, Ateş E, Kılıçkesmez Ö. Radiomics with artificial intelligence: a practical guide for beginners. Diagn Interv Radiol. 2019;25:485–495. doi: 10.5152/dir.2019.19321

6. Hatt M, Krizsan AK, Rahmim A, et al. Joint EANM/SNMMI guideline on radiomics in nuclear medicine. Eur J Nucl Med Mol Imaging. 2023;50:352–375. doi: 10.1007/s00259-022-06001-6

7. Huang EP, O’Connor JPB, McShane LM, et al. Criteria for the translation of radiomics into clinically useful tests. Nat Rev Clin Oncol. 2023;20:69–82. doi: 10.1038/s41571-022-00707-0

8. Zwanenburg A, Vallières M, Abdalah MA, et al. The Image Biomarker Standardization Initiative: Standardized Quantitative Radiomics for High-Throughput Image-based Phenotyping. Radiology. 2020;295:328–338. doi: 10.1148/radiol.2020191145

9. Orlhac F, Soussan M, Maisonobe J-A, Garcia CA, Vanderlinden B, Buvat I. Tumor Texture Analysis in 18F-FDG PET: Relationships Between Texture Parameters, Histogram Indices, Standardized Uptake Values, Metabolic Volumes, and Total Lesion Glycolysis. J. Nucl. Med. 2014;55:414–422. doi: 10.2967/jnumed.113.129858

10. Varoquaux G, Cheplygina V. Machine learning for medical imaging: methodological failures and recommendations for the future. npj Digit Med. 2022;5:1–8. doi: 10.1038/s41746-022-00592-y

11. Uno H, Cai T, Pencina MJ, D’Agostino RB, Wei LJ. On the C-statistics for evaluating overall adequacy of risk prediction procedures with censored survival data. Stat Med. 2011;30:1105–1117. doi: 10.1002/sim.4154

12. Hernán MA, Hernández-Díaz S, Werler MM, Mitchell AA. Causal Knowledge as a Prerequisite for Confounding Evaluation: An Application to Birth Defects Epidemiology. Am. J. Epidemiol. 2002;155:176–184. doi: 10.1093/aje/155.2.176

13. Benjamini Y, Krieger AM, Yekutieli D. Adaptive linear step-up procedures that control the false discovery rate. Biometrika. 2006;93:491–507. doi: 10.1093/biomet/93.3.491

14. Horvath S. Weighted Network Analysis: Applications in Genomics and Systems Biology. Springer Science & Business Media; 2011.

15. Pedregosa F, Varoquaux G, Gramfort A, et al. Scikit-learn: Machine Learning in Python. J Mach Learn Res. 2011;12:2825–2830.

16. Pölsterl S. scikit-survival: a library for time-to-event analysis built on top of scikit-learn. J Mach Learn Res. 2020;21:212:8747-212:8752.

17. Thieblemont C, Tilly H, Gomes da Silva M, et al. Lenalidomide Maintenance Compared With Placebo in Responding Elderly Patients With Diffuse Large B-Cell Lymphoma Treated With First-Line Rituximab Plus Cyclophosphamide, Doxorubicin, Vincristine, and Prednisone. J Clin Oncol. 2017;35:2473–2481. doi: 10.1200/JCO.2017.72.6984

18. Casasnovas R-O, Ysebaert L, Thieblemont C, et al. FDG-PET–driven consolidation strategy in diffuse large B-cell lymphoma: final results of a randomized phase 2 study. Blood. 2017;130:1315–1326. doi: 10.1182/blood-2017-02-766691

19. Cottereau A-S, Meignan M, Nioche C, et al. Risk stratification in diffuse large B-cell lymphoma using lesion dissemination and metabolic tumor burden calculated from baseline PET/CT. Ann. Oncol. 2021;32:404–411. doi: 10.1016/j.annonc.2020.11.019

20. Cottereau A-S, Nioche C, Dirand A-S, et al. 18F-FDG PET Dissemination Features in Diffuse Large B-Cell Lymphoma Are Predictive of Outcome. J. Nucl. Med. 2020;61:40–45. doi: 10.2967/jnumed.119.229450

21. Sasanelli M, Meignan M, Haioun C, et al. Pretherapy metabolic tumour volume is an independent predictor of outcome in patients with diffuse large B-cell lymphoma. Eur J Nucl Med Mol Imaging. 2014;41:2017–2022. doi: 10.1007/s00259-014-2822-7

22. van Griethuysen JJM, Fedorov A, Parmar C, et al. Computational Radiomics System to Decode the Radiographic Phenotype. Cancer Res. 2017;77:e104–e107. doi: 10.1158/0008-5472.CAN-17-0339

23. Møller MB, Christensen BE, Pedersen NT. Prognosis of localized diffuse large bLcell lymphoma in younger patients. Cancer. 2003;98(3):516–21. doi:10.1002/cncr.11497

24. Peduzzi P, Concato J, Kemper E, Holford TR, Feinstein AR. A simulation study of the number of events per variable in logistic regression analysis. J. Clin. Epidemiol. 1996;49:1373–1379. doi: 10.1016/S0895-4356(96)00236-3

